# Characteristics of COVID-19 recurrence: a systematic review and meta-analysis

**DOI:** 10.1101/2020.09.05.20189134

**Authors:** Tung Hoang

**Author notes:** **Correspondence:** Tung Hoang, MPH, BPharm., Institute of Research and Development, Duy Tan University, Da Nang 550000, Vietnam, Faculty of Pharmacy, Duy Tan University, Da Nang 550000, Vietnam.

## Abstract

**Background:** Previous studies reported the recurrence of coronavirus disease 2019 (COVID-19) among discharge patients. This study aimed to examine the characteristic of COVID-19 recurrence cases by performing a systematic review and meta-analysis.

**Methods:** A systematic search was performed in PubMed and Embase and gray literature up to September 17, 2020. A random-effects model was applied to obtain the pooled prevalence of disease recurrence among recovered patients and the prevalence of subjects underlying comorbidity among recurrence cases. The other characteristics were calculated based on the summary data of individual studies.

**Results:** A total of 41 studies were included in the final analysis, we have described the epidemiological characteristics of COVID-19 recurrence cases. Of 3,644 patients recovering from COVID-19 and being discharged, an estimate of 15% (95% CI, 12% to 19%) patients was re-positive with SARS-CoV-2 during the follow-up. This proportion was 14% (95% CI, 11% to 17%) for China and 31% (95% CI, 26% to 37%) for Korea. Among recurrence cases, it was estimated 39% (95% CI, 31% to 48%) subjects underlying at least one comorbidity. The estimates for times from disease onset to admission, from admission to discharge, and from discharge to RNA positive conversion were 4.8, 16.4, and 10.4 days, respectively.

**Conclusion:** This study summarized up-to-date evidence from case reports, case series, and observational studies for the characteristic of COVID-19 recurrence cases after discharge. It is recommended to pay attention to follow-up patients after discharge, even if they have been in quarantine.

## Introduction

Since December 2019, the world has been experiencing a public health crisis due to severe acute respiratory syndrome coronavirus-2 (SARS-CoV-2). As of September 01, 2020, about 26 million confirmed cases and 0.8 million deaths were reported from 213 countries and territories ^1^. Several nationwide studies retrospectively investigated clinical features and the epidemiological characteristics of patients infected with SARS-CoV-2 ^2-4^. Particularly, aging and underlying chronic diseases were reported to much contribute to the severity of coronavirus disease 2019 (COVID-19) ^5,6^. However, patients with COVID-19 were generally less severe than SARS and Middle East respiratory syndrome (MERS), with the fatality rate of 9.6%, 34.3%, and 6.6% for SARS, MERS, and COVID-19, respectively ^7^. Recently, it has been reported that SARS-CoV-2 RNA shedding duration could prolong up to 83 days ^8^. In addition, the repositive SARS-CoV-2 RNA test has been observed among patients who had been discharged from health care units and received regular follow-up ^8^. Therefore, this systematic review and meta-analysis was conducted to examine the prevalence of the RNA repositive test for SARS-CoV-2 among recovered patients, the prevalence of subjects underlying comorbidity among recurrence cases, in addition to times from disease onset to hospital admission, from admission to hospital discharge, and from discharge to positive RNA conversion.

## Methods

An electronic search of PubMed and Embase was conducted for English language studies published from the inception until September 17, 2020. The keywords for searching were as follows: “(COVID-19 OR SARS-CoV-2) AND (recurrence OR recurrences OR reinfection OR re-infection)”. Additionally, hand searching for related reports of the Centers for Disease Controls and bibliography of relevant studies was performed to obtain relevant information. For each study, the following information was extracted: first author’s name, country, study type, number of recurrence cases and discharged patients, the sample used for reverse transcription polymerase chain reaction (RT-PCR), mean or median age (years), number of males, females, and cases underlying any chronic diseases (including chronic obstructive pulmonary disease, cardiovascular disease, hypertension, diabetes, liver or kidney disease, and cancer), times from disease onset to admission, from admission to discharge, and from discharge to positive conversion (days). In this study, heterogeneity was quantified by the I^2^ statistics, in which I^2^>50% was defined as potential heterogeneity ^9^. Given data are from different populations of various characteristics, a random-effects model was used to calculate the pooled effect size and its 95% confidence interval (CI) when the evidence from at least two individual studies was available ^10^. All the statistical analyses were performed using STATA 14.0 software.

## Results

The study selection process is presented in **Figure 1**. Initial 550 records were retrieved through PubMed (N=239) and Embase (N=311) and additional 1 gray literature through hand searching was identified. Among records after removing duplicates and non-English publications (N=128), 423 studies were potentially relevant through reviewing titles and abstracts. After reviewing full-text articles, 15 studies were excluded because they reported overlapping cases (N=6) or irrelevant population (N=3), there was no information for outcomes of interest (N=4), and they were studies of mechanisms or modeling (N=2). The remaining 41 studies were therefore eligible for the final analysis ^11-51^.

**Figure 1.**
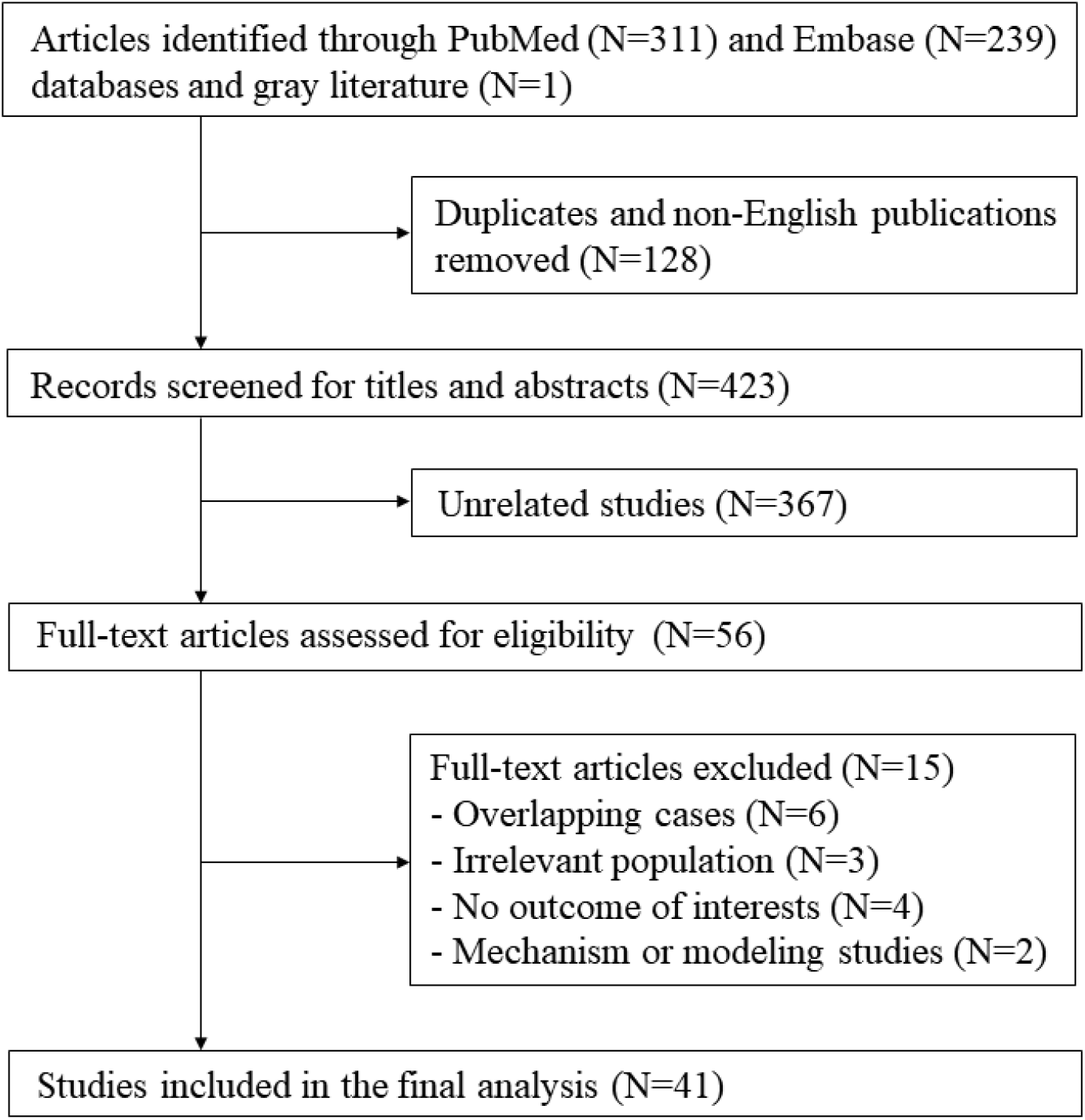
Flowchart of study selection

A detailed description of extracted data of included studies is shown in **Table 1**. Thirty-eight studies reported 466 recurrence cases from China (N=33, 435 cases), Korea (N=1, 83 cases), Iran (N=1, 1 case), Brunei (N=1, 21 cases), Italy (N=2, 3 cases), France (N=1, 11 cases), Brazil (N=1, 1 case), and US (N=1, 1 case). The study design included case reports (N=14), case series (N=6), and observational studies (N=21).

**Table 1.** Summary of studies reporting recurrence of COVID-19 cases after discharge

| Study | Country | Study type | No. of recurrence cases | No. of discharged patients | Sample for testing | Age (years) | Male/female | No. of cases underlying comorbidity | Times from onset to admission (days) | Times from admission to discharge (days) | Times from discharge to positive conversion (days) |
| --- | --- | --- | --- | --- | --- | --- | --- | --- | --- | --- | --- |
| Alonso FOM | Brazil | Case report | 1 |  | Respiratory swab | 26 | 1/0 |  |  |  | 34 |
| An J | China | Observational | 38 | 242 | Nasal and anal swab | 32.8 | 16/22 |  |  |  |  |
| Batisse D | France | Case series | 11 |  | Naso-pharyngeal swabs | 55 | 6/5 | 7 |  |  |  |
| Bongiovanni M | Italy | Case series | 2 |  | Nasopharyngeal swab |  | 0/2 | 2 |  |  |  |
| Cao H | China | Observational | 8 | 108 | Deep nasal cavity or throat swab | 54.4 | 3/5 | 0 |  |  | 16.3 |
| Chen D | China | Case report | 1 |  | Oropharyngeal swab | 46 | 1/0 |  | 8 |  |  |
| Chen J | China | Observational | 81 | 1087 | Throat swab | 62 | 30/51 | 29 |  | 12 | 9 |
| Chen Y | China | Observational | 4 | 17 | Oropharyngeal, nasopharyngeal, and anal swab | 32 | 2/2 |  |  | 18.25 | 11.25 |
| Duggan NM | US | Case report | 1 |  |  | 82 | 1/0 | 1 | 7 | 39 | 10 |
| Fu W | China | Case series | 3 |  | Nasopharyngeal swab | 48 | 1/2 |  |  | 12 | 9.3 |
| Gao G | China | Case report | 1 |  |  | 70 | 1/0 | 1 | 5 | 15 | 12 |
| Geling T | China | Case report | 1 |  | Pharyngeal swab | 24 | 1/0 | 0 |  | 10 | 8 |
| He F | China | Case report | 1 |  | Throat swab | 39 | 0/1 |  | 10 | 13 | 8 |
| Hu R | China | Observational | 11 | 69 | Nasopharyngeal swab | 27 | 7/4 | 3 |  | 10 | 14 |
| Huang J | China | Observational | 69 | 414 | Nasopharyngeal and anal swab |  | 28/41 | 22 | 3 | 20 | 11 |
| KCDC | Korea | Observational | 83 | 269 |  |  | 28/41 |  |  |  | 14.3 |
| Li J | China | Case report | 1 |  | Nasopharyngeal and oropharyngeal samples | 71 | 0/1 |  | 14 |  |  |
| Li XJ | China | Case report | 1 |  |  | 41 | 1/0 |  | 19 | 9 | 19 |
| Li Y | China | Observational | 6 | 13 | Oral swabs, nasal swabs, sputum, blood, faeces, urine, vaginal secretions, and milk | 51.3 | 3/3 | 3 |  |  | 10.2 |
| Liang C | China | Observational | 11 | 22 | Throat swab |  |  |  |  |  |  |
| Liu T | China | Observational | 11 | 150 | Throat swab | 49 | 6/5 |  |  |  |  |
| Loconsole D | Italy | Case report | 1 |  | Nasopharyngeal swab | 48 | 1/0 | 0 |  | 15 | 30 |
| Luo A | China | Case report | 1 |  | Throat swab | 58 | 0/1 |  | 7 | 15 | 22 |
| Mardani M | Iran | Case report | 1 |  | Nasopharyngeal swab | 64 | 0/1 |  |  |  |  |
| Peng J | China | Case series | 7 |  | Throat swab |  | 4/3 |  |  | 16.7 | 10.1 |
| Qiao XM | China | Observational | 1 | 15 | Nasopharyngeal and throat swab | 30 | 0/1 |  |  | 14 | 15 |
| Qu YM | China | Case report | 1 |  | Throat swab and sputum | 49 | 1/0 |  | 4 |  |  |
| Tian M | China | Observational | 20 | 147 | Pharyngeal swabs | 37.15 | 11/9 | 7 | 2.5 | 18.65 | 17.25 |
| Wang P | China | Case report | 1 |  | Throat swab | 33 | 1/0 |  | 8 | 21 | 15 |
| Wang X | China | Observational | 8 | 131 |  | 48.75 | 4/4 | 0 |  |  | 11.375 |
| Wong J | Brunei | Observational | 21 | 106 | Nasopharyngeal swab | 43.1 | 12/9 |  |  | 17 | 13 |
| Xiao AT | China | Observational | 15 | 70 | Throat swab, deep nasal cavity swab | 64 | 9/6 |  |  |  |  |
| Xing Y | China | Case series | 2 |  | Throat swab and stool tests |  | 1/1 |  | 6 | 15.5 | 6.5 |
| Ye G | China | Observational | 5 | 55 | Throat swab | 32.4 | 2/3 | 0 |  |  | 10.6 |
| Yuan B | China | Observational | 20 | 182 | Nasopharyngeal swab or anal swab | 39.9 | 7/13 | 6 | 5.1 | 20.8 | 9.45 |
| Yuan J | China | Observational | 25 | 172 | Cloacal swab and nasopharyngeal swab | 28 | 8/17 |  |  | 15.36 | 5.23 |
| Zhang B | China | Case series | 7 |  | Throat and rectal swab | 22.4 | 6/1 |  |  | 15.4 | 9.7 |
| Zheng KI | China | Observational | 3 | 20 | Salivary and fecal |  |  |  |  |  | 7 |
| Zhou X | China | Case report | 1 |  | Oropharyngeal swab | 40 | 1/0 |  | 6 | 16 | 7 |

| **Study** | **Country** | **Study type** | **No. of recurrence cases** | **No. of discharged patients** | **Sample for testing** | **Age (years)** | **Male/ female** | **No. of cases underlying comorbidity** | **Times from onset to admission (days)** | **Times from admission to discharge (days)** | **Times from discharge to positive conversion (days)** |
| --- | --- | --- | --- | --- | --- | --- | --- | --- | --- | --- | --- |
| Zhu H | China | Observational | 17 | 98 | Sputum and pharyngeal swab | 54 | 5/12 |  |  |  | 4 |
| Zou Y | China | Observational | 53 | 257 | Throat swabs | 62.19 | 23/30 | 29 |  |  | 4.6 |

The calculation of the epidemiological characteristics of COVID-19 recurrence cases is presented in **Table 2**. Data for age were provided from 34 studies for 379 recurrence cases, with a mean age of 41.7 years. Among 542 recurrence cases from 39 studies, 233 cases were males, which accounted for 43%. Times from disease onset to admission, from admission to discharge, and from discharge to RNA positive conversion were available for 52, 276, 464 cases from 13, 22, and 31 studies, respectively. The estimates for times from disease onset to admission, from admission to discharge, and from discharge to RNA positive conversion were 4.8, 16.4, and 10.4 days, respectively.

**Table 2.** Epidemiological characteristics of COVID-19 recurrence cases

| **Characteristic** | **No. studies** | **No. of recurrence cases** | **Result** |
| --- | --- | --- | --- |
| Age (years) | 34 | 379 | 41.7 |
| Male (no., %) | 39 | 233 | 542 (43%) |
| Times from onset to admission (days) | 13 | 52 | 4.8 |
| Times from admission to discharge (days) | 22 | 276 | 16.4 |
| Times from discharge to positive conversion (days) | 31 | 464 | 10.4 |

The prevalence of COVID-19 recurrence cases after discharge was calculated from data of 21 observational studies (**Figure 2**). Among 3,644 discharged patients, the RT-PCR test turned to be positive in 406 Chinese, 83 Korean, and 21 Bruneian subjects. Overall, the prevalence of recurrence cases was 15% (95% CI, 12% to 19%). Substantial heterogeneity among studies was observed, with I^2^ of 86.32%. In the subgroup analysis by population, the prevalence was reported to be 14% (95% CI, 11% to 17%) for China, 31% (95% CI, 26% to 37%) for Korea, and 20% (95% CI, 13% to 28%) for Brunei.

**Figure 2.**
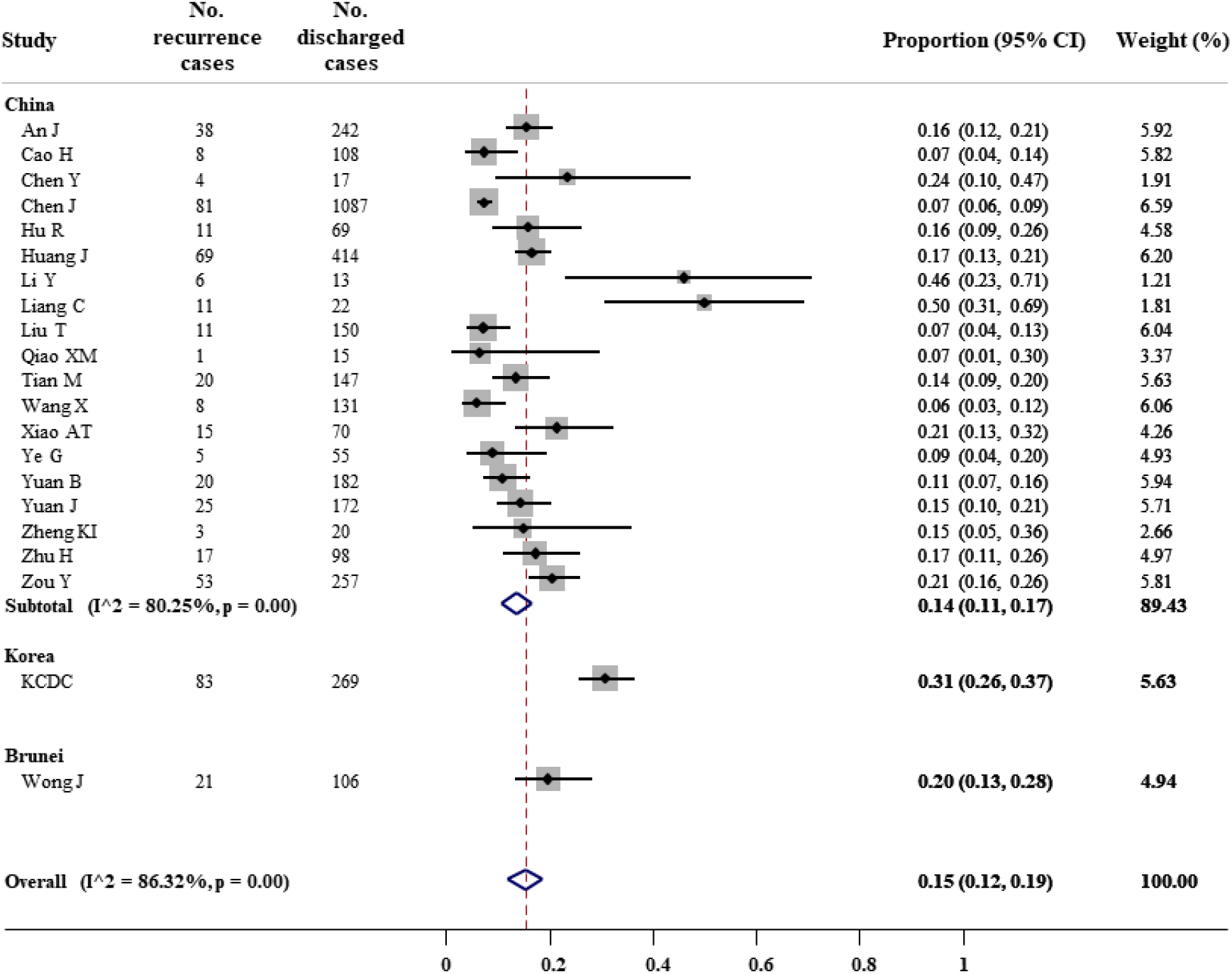
Forest plot for meta-analysis of COVID-19 recurrence prevalence

Furthermore, it was reported 106 subjects underlying comorbidity among a total of 271 recurrence cases, which accounted for 39% (95% CI, 31% to 48%) (**Figure 3**). There was no evidence of heterogeneity (I^2^=42.08%). Subgroup analysis showed the proportion of 64% (95% CI, 35% to 85%) for France cases and 38% (30% to 45%) for Chinese cases.

**Figure 3.**
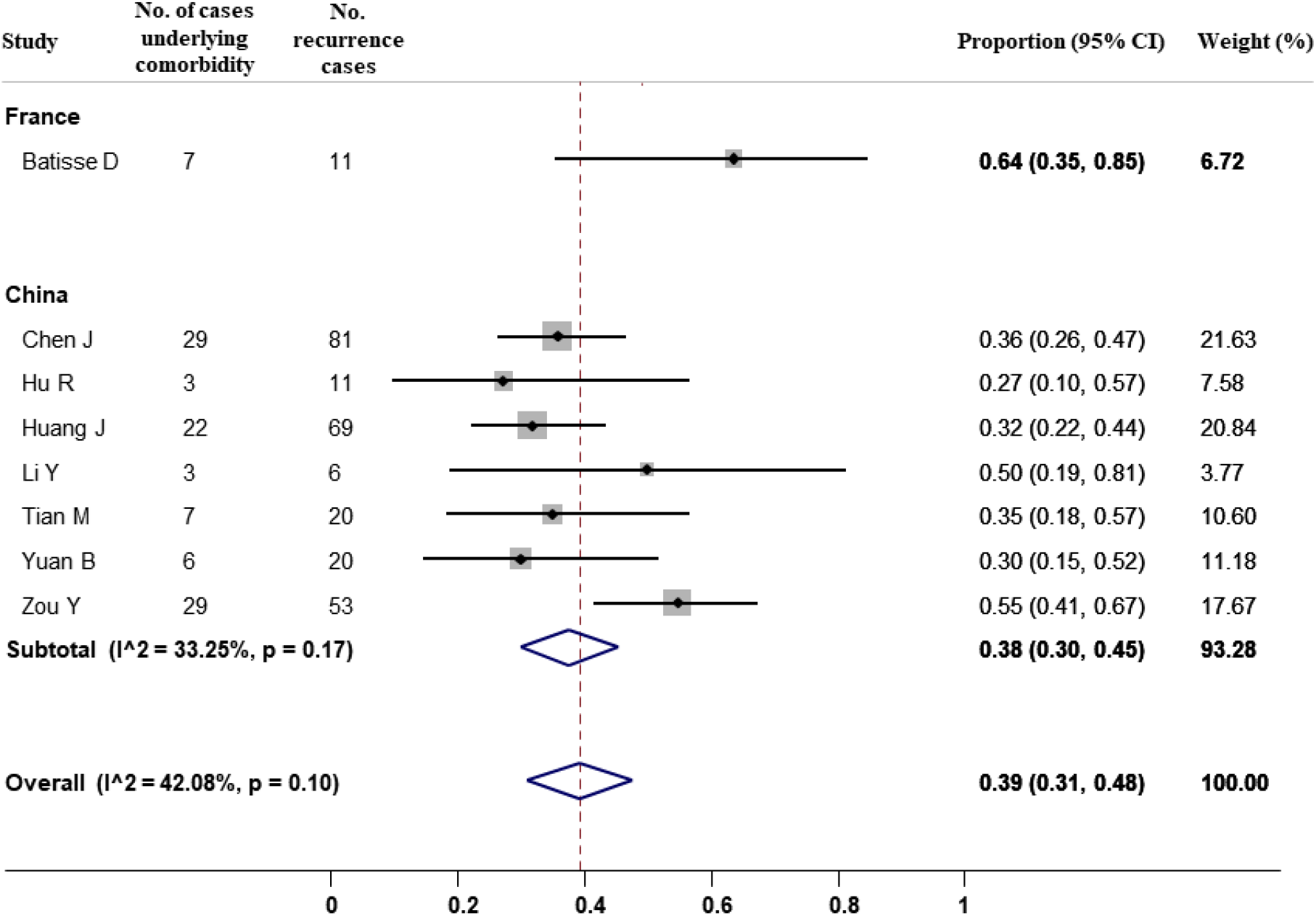
Forest plot for meta-analysis of comorbidity among COVID-19 recurrence cases

## Discussion

In this systematic review and meta-analysis of 41 studies, we have described the epidemiological characteristics of COVID-19 recurrence cases. Of 3,644 patients recovering from COVID-19 and being discharged, an estimate of 15% (95% CI, 12% to 19%) of patients was repositive with SARS-CoV-2 during the follow-up. This proportion was 14% (95% CI, 11% to 17%) for China, 31% (95% CI, 26% to 37%) for Korea, and 20% (95% Ci, 13% to 28%) for Brunei. Among recurrence cases, it was estimated 39% (95% CI, 31% to 48%) subjects underlying at least one comorbidity.

According to the guidelines of the World Health Organization, a patient can be discharged from the hospital after two consecutive negative results in a clinically recovered patient at least 24 hours apart ^52^. However, the discharge criteria for confirmed COVID-19 cases are additionally required according to different countries ^53^. The determination of recurrence cases can be caused by false negatives, which ranged from 2% to 29% according to a meta-analysis of 957 hospitalized patients ^54^. The reason for false negatives can be due to the source of specimens, sampling procedure, and the sensitivity and specificity of the test kit ^8^. In a preprint study of 213 Chinese patients, a total of 205 throat swabs, 490 nasal swabs, and 142 sputum samples were collected, and the false-negative rates were reported of 40%, 27%, and 11% for the throat, nasal, and sputum samples, respectively ^55^. Due to the lack of individual data, we were not able to examine the prevalence of recurrence cases in the subgroup analysis by types of specimens.

Furthermore, it may require considering prolonged SARS-CoV-2 shedding in asymptomatic or mild cases and recurrence of viral shedding ^56^. Data from 68 patients revealed a significantly longer duration of viral shedding from sputum specimens (34 days) than nasopharyngeal swabs (19 days) ^57^. Consistent findings were reported in an asymptomatic case with viral detection positive in stool but negative in nasopharyngeal swab lasts for 42 days ^58^. Similarly, the positive rate of the SARS-CoV-2 RNA test was shown to be highest for the sputum sample (100%), followed by nasal swab (75%), oral swab (40%), and stool specimen (38%) ^59^. Nevertheless, although the RT-PCT results of discharge patients were possible to turn positive, it is necessary to distinguish between reactivation and reinfection cases ^8^.

Factors related to the recurrence of COVID-19 remain unclear because of inconsistent findings. Although disease severity may be associated with the worse immune response, An J *et al*. reported the lower recurrence rate among subjects with severe or moderate disease at baseline than those with mild disease (odds ratio [OR]=0.23, 95% CI=0.10-0.53) ^12^. However, the proportion in subjects with severe disease did not differ in those with moderate or mild disease (OR=1.06, 95% CI=0.57-1.96) ^17^. Also, while subjects underlying diseases such as hypertension and diabetes are more likely to be susceptible with disease infection and severity ^60^, the recurrence proportion was not significantly different between comorbidity carriers and noncarriers, in Chen *et al*.’s study (OR=0.71, 95%=0.42-1.20 for hypertension and OR=0.85, 95% CI=0.42-1.75 for diabetes) ^17^ and Huang *et al*.’s study (OR=0.98, 95%=0.52-1.87 for hypertension and OR=0.46, 95% CI=0.14-1.55 for diabetes) ^25^.

This study summarized up-to-date evidence from case reports, case series, and observational studies for the characteristic of COVID-19 recurrence cases after discharge. However, several limitations need to be mentioned. First, 80% of the included studies (33/41) with 78% recurrence cases (435/556) come from the Chinese population, which may reduce the availability to generalize the pooled estimates into other populations. Second, heterogeneity for the prevalence of recurrence cases was substantially presented among studies. The different characteristics, discharge criteria, and the test samples used among study populations included in this meta-analysis may have contributed to the heterogeneity. Last, all the estimates in the current study are based on aggregate data from published articles. Failure to obtain individual patient data may lead to bias due to the lack of full exploration and adjustment for patient characteristics ^61^.

In summary, an estimate of 15% of COVID-19 patients was repositive after discharge. Among them, 39% of subjects were underlying comorbidity. It is recommended to pay attention to follow-up patients after discharge by closely monitoring their RT-PCR results, even if they have been in quarantine for 14 days. Further studies are needed to determine factors associated with positive RT-PCR in COVID-19 patients after discharge.

## Data Availability

Data for all the analyses are available in Table 1.

## Data availability statement

Data for all the analyses are available in Table 1.

## Author’s contributions

TH designed the outline of the work, analyzed the data, and wrote the

